# Serum-derived extracellular vesicles as biological indicator of mobility resilience in older adults

**DOI:** 10.1101/2025.09.08.25335349

**Authors:** Nicholas F. Fitz, Ashish Kumar, Yixin Su, Mitu Sharma, Sangeeta Singh, Radosveta Koldamova, Iliya Lefterov, Amrita Sahu, Fabrisia Ambrosio, Caterina Rosano, Gagan Deep

## Abstract

The decline in mobility with aging is a major health concern, associated with a high risk for disability. Despite the widespread prevalence of gait slowing in elderly adults, this issue has not been adequately addressed. The central nervous system and skeletal muscle system are key regulators of gait speed. However, direct molecular communication along the brain-muscle axis and the role of these interactions in mobility resilience remain poorly studied. Recently, extracellular vesicles (EV), membrane bound vesicles secreted by cells, have emerged as a key player in long distance inter-cellular communication. Nevertheless, the potential of EVs as biological predictor of mobility resilience in older adults has not been yet studied. In the present study, we used serum samples from 23 participants with gait speed >1.0 m/sec (mobility-resilient group) and 22 participants with gait <1.0 m/sec (mobility non-resilient group) from the Health, Aging and Body Composition (Health ABC) study. First, total circulating serum EVs were isolated and characterized for small noncoding RNAs using un-biased small noncoding RNA sequencing. Given the central role of mitochondria in muscle energy metabolism and their emerging link to age-related physical decline, next, muscle-derived EVs (MDE) were isolated and characterized for specific mitochondrial markers (TOM20, mtCox2, PDH, and VDAC) by flow cytometry, the expression of a panel of 13 miRNAs related to mitochondrial function by RT-PCR, and PPAR-γ expression by ELISA. The results showed differential enrichment of various miRNAs, circRNAs, and mitochondrial proteins in total EVs and/or MDE between mobility-resilient and non-resilient groups, highlighting their potential as non-invasive biomarkers for mobility outcomes. Overall, the findings from the present study suggest a role for serum EVs in mediating molecular communication related to functional aging phenotypes and underscores the potential of EV biomarkers in modulating mobility and promoting healthy aging.

## Introduction

Slow walking gait with aging is a major health concern linked to increased risk of frailty and dementia, loss of independence, increased health care use, and reduced survival rates^1,2^. With aging, elderly adults (>70 years) can develop slow gait in the absence of a clear diagnosis or cause. In fact, we showed that ∼80% of older adults with more than one locomotor risk factor (e.g., muscle weakness, joint pain, etc.) have mobility decline and considered mobility non-resilient (gait speed < 1.0 m/second), which is associated with high risk for disability^3^. While other individuals with similar locomotor risk factors maintain mobility resilient (gait speed > 1.0 m/second) with aging. Despite the high prevalence of gait slowing in the community, this decline in mobility and activity is inadequately addressed and the underlying mechanisms are poorly understood. Although slow walking is widely recognized as a multisystemic phenomenon, alterations in the central nervous system (CNS) and skeletal muscle system (SMS) have emerged as significant predictors of gait speed and point to a brain-muscle axis that plays a critical role in mobility resilient. These systems exert critical modulatory and interactive influences on mobility performance, independent of the severity of underlying locomotor risk factors. The coordination, integrity, and functional capacity of the CNS and SMS play a pivotal role in maintaining faster gait and overall mobility, even amidst age-related declines in other physiological systems. Notably, both the CNS and SMS remain adaptable later in life and influence each other through neural pathways and circulating biological factors, making them promising targets for interventions aimed at sustaining mobility in aging populations

While there are direct nerve connections between the brain and skeletal muscles via efferent motor axons and afferent sensory axons, other pathways are also important in the brain and muscle crosstalk. One potential candidate to coordinate brain and muscle communication is extracellular vesicles (EVs). EVs are lipid membrane-delimited nanoscale vesicles secreted by almost all cells into the interstitial space and contain specific cargos, including nucleic acids, proteins, metabolites, and lipids^4–8^. EVs are heterogeneous in size and can be categorized as small EVs (∼50-<200 nm), large EVs (200-1000 nm), and very large EVs (1-10 µm)^9^. Due to the nano size of small EVs, they can travel long-distance via circulatory system and could play a critical role in exchanging molecular information between remote sites in the body. EVs cargoes, taken up by target cells, can regulate their physiological functions or pathological processes in recipient cells. Importantly, the content of EVs, at the core and/or the surface allows identification of organ specificity, including brain or muscle specific EVs^10^. In multiple studies, we have reported the utility of EVs for examining the pathophysiological state of different cells and tissues, including brain cells and muscle tissues^6–8,10–13^. Notably EVs, regardless of their origin, are enriched with extracellular RNAs of many different types, particularly small noncoding RNAs (ncRNAs). The most frequently reported types of small ncRNAs carried by small EVs include: miRNAs (MicroRNAs), tRNAs (Transfer RNAs), piRNAs (PIWI-interacting RNAs), snRNAs (Small nuclear RNAs), snoRNA (Small nucleolar RNAs), and circRNAs (Circular RNAs). While the exact functions of various small ncRNAs are not fully known, studies have shown their ability to regulate gene expression at the level of posttranscriptional messenger RNA (mRNA) processing^14–16^. For example, miRNAs bind to a specific sequence of target mRNAs to induce translation repression through either mRNA silencing or degradation. Initially it was believed that miRNAs bind the 3’ untranslated region, but more recently, binding sites have been identified in the 5′ untranslated region, coding sequence, and gene promoter regions^17^. In contrast, circRNAs can act as microRNA sponges, inhibiting miRNA modulation of gene expression^18^. Therefore, circulating EVs carry small ncRNAs which can act either as activators or suppressors of gene expression in a complex relationship^19,20^.

Notably, it has been proposed that physical exercise and the skeletal muscle secretome reduces age-related brain atrophy, oxidative stress, neuroinflammation, and the progression of neurodegenerative diseases while supporting cognitive function and mitochondrial biogenesis^5,21–25^. Circulating factors such as myokines, growth factors, hormones, and cytokines have been proposed as mediators of this brain-muscle communication^25,26^. However, their ability to influence central processes may be constrained by limited permeability across the blood-brain barrier (BBB). We and others showed that EVs are key messengers of paracrine exercise signals^27–30^. EVs possess the unique capacity to traverse the BBB, enabling bidirectional signaling between the brain and peripheral tissues. This is achieved through the delivery of bioactive cargo, including small non-coding RNAs, which may play a critical role in coordinating systemic physiological responses^31–34^. These vesicles may carry both neuroprotective and myoprotective factors capable of crossing the BBB, thereby linking central and peripheral health in aging and disease contexts^5,11,21^. Also, muscle-derived EVs (MDE) have been shown to have significant role in mediating the crosstalk between skeletal muscles and other organs/ tissues^35^. This communication has also been suggested by a previous study by Aswad et al., which demonstrated that fluorescently labeled skeletal muscle derived EVs from mice can be taken up by various tissues including brain^36^. Moreover, myoblasts were also identified to release EVs, and during differentiation, these EVs contained muscle growth factors related to muscle development^37–39^. Furthermore, we have shown the utility of neuromuscular electrical stimulation (NMES) to promote the release of circulating EVs that have both myo- and neuro-protective effects, which increased the ability to repair injured skeletal muscle in aged mice^40^. We also observed higher levels of neuroprotective molecules in EVs, e.g. such as BDNF (Brain-derived neurotrophic factor), following muscle contractile activity supporting that circulating EVs may serve as key mediators in this axis by transporting neuroprotective and myoprotective factors to target organs^40^.

We hypothesized that brain and skeletal muscle crosstalk not only regulates the functions of each system but ultimately influences gait speed. We further hypothesized that circulating EV cargos contain molecular messengers that are important in modulating mechanisms underlying the brain-muscle axis. While there is abundant data that the CNS and SMS have beneficial effects on each other, and that enhancing one can lead to improvements in the other and mobility, this evidence is primarily from preclinical models and studies of each system in isolation. Analysis of the circulatory EVs for their cargos, particularly ncRNA could be valuable due to their regulatory function in mediating gene expression and cellular signaling and can provide insight into the possible molecular signals and mechanisms facilitating communication between these two distant organs. Moreover, since mitochondria play a central role in muscle health, mobility, and gait speed, particularly through their functions in energy production, cellular maintenance, and adaptation to stress, mitochondrial proteins were studied in MDE to better understand the molecular mechanisms underlying age-related declines in physical performance. We first conducted a comprehensive profiling of circulating total EVs for small ncRNA, followed by MDE cargos for mitochondrial markers using the serum samples of age-matched older individuals, classified as mobility resilient or non-resilient. These interactions have not been previously tested in the context of slow gait speed, aging, or accounting for other locomotor risk factors. We identified unique small ncRNAs in EVs isolated from the serum of mobility resilient compared with non-resilient individuals. Furthermore, many of the small ncRNAs have been shown to be important in regulating genes involved in brain health and metabolism. Further, the mitochondrial markers showed higher levels in MDE from the mobility resilient group. We also show correlations of these mitochondrial markers with mobility phenotypes in the two cohorts. These results can help inform future studies to determine the importance of EV cargos in pathological mechanisms of the CNS and SMS and potentially identify biomarkers which would allow for detection of early changes in mobility or monitor interventions to help improve mobility among elderly individuals.

## Materials and Methods

### Study population

The Health, Aging and Body Composition (Health ABC) Study recruited 3,075 men and women aged 70-79 living in Memphis, Tennessee, or Pittsburgh, Pennsylvania, to study changes in body composition, functional limitation, disability, and mortality^41^. Participants were eligible if they were free of difficulty when walking ¼ of a mile and free of difficulty climbing 10 stairs. All participants (60% women, 40% African American) had active follow up starting in 1997 – 1998 (Year 1, or baseline of the study) and ending in 2011. The Health ABC study has archived blood samples from 2,893 older adults concurrent with muscle imaging, mobility and cognitive measures as well as extensive health characterization. Of these 2,893, we identified those with impairment in at least one system important for locomotion, as previously described, including: joint pain, peripheral arterial disease, cerebrovascular and cardiorespiratory diseases or conditions^3^. Mobility resilient was defined as at least one locomotor risk factor (pain, high BMI, low forced expiratory volume, peripheral artery disease) and a gait speed >1.0m/sec, while non-resilient individuals were defined as one locomotor risk factor and gait speed <1.0m/sec. Among these, we randomly selected 23 participants with gait speed ≥1.0 m/sec (resilient) and 22 with gait <1.0m/sec (non-resilient), for a total of 45 participants. Blood samples from 21 of these 45 participants (11 resilient, 10 non-resilient) were assessed at Wake Forest School of Medicine (WFSM, Site 1) and the remaining 24 (n=12 each of resilient and non-resilient) were assessed at the University of Pittsburgh (UPitt, Site 2). Blood samples were collected at the year 2 study visit (1998-99) in the morning after overnight fasting for at least eight hours. We used serum samples that had not been previously thawed and were stored at −80°C from the time of collection.

Measures of interest included age, sex and race by self-report, and cognitive function, muscle strength and comorbidities measured as previously described^42^. Briefly, the Digit Symbol Substitution Test (DSST), tested executive function and information processing skills; it ranges: 0-100, with higher scores indicating better executive functioning^43^. The modified mini-mental exam (3MS) is a validated cognitive screening tool, widely used as a marker of cognitive change and incident dementia in older adult populations^44^; it ranges 0-90, with higher scores indicating better function^45^. Muscle strength was measured as right knee extension strength with an isokinetic dynamometer (Kin-Com dynamometer, 125 AP; Chattanooga, TN); the left leg was tested for those with joint replacement or knee pain on the right side. Other contraindications to strength testing included: systolic blood pressure greater than or equal to 200 mmHg, diastolic blood pressure greater than or equal to 110 mmHg, history of cerebral aneurysm, cerebral bleeding, bilateral total knee replacement, or severe bilateral knee pain (12.7% of original cohort). The maximum muscle torque (Newton meters) was calculated from the average of three reproducible and acceptable trials from a maximum of six.

The presence of comorbidities was assessed via self-reported questionnaires, medications listed, or Health Care Finance Administration diagnosis. Total weekly energy expenditure was calculated as kilocalories per kilogram per week based on self-reported physical activity. Body mass index was computed based on measurements of weight divided by height. All investigators involved with EV assessments were blinded to group assignment.

### Total EV isolation from serum for small RNA sequencing

EVs from serum were isolated using Plasma/Serum Exosome Purification Mini Kit (Norgen Biotech Corp.) according to the manufacturer’s instruction. 400 µL serum was used for EV isolation in which 3.6 mL particle free water was added. To each serum sample, 100 µL of ExoC buffer was added followed by 200 µL of Slurry E and incubated at room temperature for 5 min. Following incubation, the samples were centrifuged for 2 min at 2,000 RPM, the supernatant was removed and 200 µL ExoR buffer added to the slurry pellet for EV release. After incubation for 5 min, samples were centrifuged for 2 min at 500 RPM and supernatant transferred to a mini filter spin column assembly and centrifuged for 1 min at 6000 RPM to isolate the EVs in the flowthrough.

### Isolation of muscle-derived EVs (MDE)

Isolation of MDE was performed from total EVs using α-sarcoglycan (SGCA) marker, following the recently published method^6^. Briefly, 1,500 µg of total EVs were incubated with 8 µg of biotin-tagged SGCA (orb453551, Biorybt) antibody overnight at 4°C with continuous mixing. Thereafter, streptavidin-tagged magnetic beads were added for 2 h incubation at room temperature with continuous mixing. Beads were washed 3 times with 0.1% Tween-20 in Tris-buffered saline and MDE were eluted by adding 200 µL of IgG elution buffer (21004; Thermo Fisher Scientific). Beads were magnetically removed, and the supernatant containing the MDE were collected in tubes containing 20 µL of 1 M tris buffer (pH 9) to neutralize the pH.

### EV characterization

For EV characterization, 1 ml of diluted EV sample (1:500) was loaded on a NS300 Nanosight instrument (Malvern Instruments) at a rate of 400 μL/min. The instrument was calibrated with 100 nm latex beads diluted in particle free PBS prior to sample analysis. Before and after the analysis of each sample, particle free PBS was used to wash the instrument. Sample analysis consisted of 3 separate measurements of 30 seconds each.

Nanoparticle Tracking Analysis (NTA) software, version 3.4, was used to visualize and measure each sample to determine particle size and concentration. EV samples were further characterized by Western blotting and transmission electron microscopy (TEM). For Western blotting, EV proteins were resolved on 4–12% Bis-Tris gels (ThermoFisher Scientific) and transferred onto nitrocellulose membranes (IB23001, Thermo Fisher Scientific, iBlot2 Gel system). These membranes were probed with anti-Calnexin (abc22595, 1:2000 dilution, secondary antibody-Santa Cruz goat anti-rabbit IgG-HRP, 1:3000); anti-Flotillin (BD Biosciences 610820, 1:1000 dilution; secondary antibody-Santa Cruz goat anti-mouse IgG-HRP, 1:3000); anti-CD63 (System Biosciences EXOAB-CD63A-1, 1:500 dilution, secondary antibody-goat anti-rabbit HRP, 1:10,000) and anti-CD81 (System Biosciences EXOAB-CD81A-1, 1:500 dilution, secondary antibody-goat anti-rabbit HRP, 1:10,000). Immunoreactive signals were visualized using enhanced chemiluminescence with the Amersham Imager 600 (GE Lifescience). TEM was performed on freshly isolated EVs as previously described^4^. Briefly, EVs were adhered to copper grids coated with 0.125% Formvar in chloroform, stained with 1% uranyl acetate in ddH₂O, and imaged immediately using a JEM 1011 transmission electron microscope.

### Flow Cytometry

To analyze the surface expression of mitochondrial markers, total EVs and MDE were incubated with fluorescently labelled, translocase of outer mitochondrial membrane 20 (TOM20), voltage-dependent anion channel (VDAC), cytochrome c oxidase subunit 2 (mtCox2), or pyruvate dehydrogenase (PDH) antibodies for overnight incubation at 4°C. Further, EVs were labelled with membrane labeling dye CellBrite 488 (Biotium, Fremont, CA, USA) for 15 min at room temperature. EVs without membrane labelling dye were used to set the gate for dye positive events (EVs). Dye labelled EVs without antibodies were used to set the gate for antibody positive events. Samples were analyzed on nano-flow cytometer (CytoFlex, Beckman Coulter, Brea, CA, USA) using the methods and machine settings described by us earlier^6–8,46^.

### RNA isolation and library preparation

Total RNA was extracted from isolated serum EVs using Exosomal RNA Isolation Kit (58000, Norgen Biotech Corp.) according to the manufacturer’s protocol. The quantification and quality of isolated RNA was evaluated with the Agilent 2100 Bioanalyzer RNA Pico assay (Agilent Technologies) following the manufacturer’s instructions. RNA was pelleted with Pellet Paint (Novagen) and reconstituted in 6 µl of nuclease free water. The NEBNext small RNA sample library preparation kit (New England Biolabs) was used according to manufacturer’s protocol with minor modifications. All reactions were with a 1:10 adapter dilution and run with 18 PCR cycles. The library products were cleaned using AMPure XP beads (Beckman Coulter). The small RNA library quality and quantity were assessed on Bioanalyzer 2100 High Sensitivity DNA chip (Agilent technologies) and sequenced at the Health Sciences Sequencing Core at UPMC Children’s Hospital of Pittsburgh, on the NextSeq 2000 machine (1×51 bp).

### Computational analysis of sequencing data

Small ncRNA library data generated from human serum and from published data of tissue atlas^47^ were aligned and quality checked by STAR (v 2.5.3a) and annotated with COMPSRA (v 1.0.3) and differential abundance determined by DESeq2 (v 1.36.0). Selected microRNA gene targets were analyzed using TargetScanHuman at cumulative weighted context score cutoff<-0.5. For circRNA, associated genes were identified by circBase database (www.circbase.org). Functional annotation clustering was performed using the Database for Annotation, Visualization and Integrated Discovery (DAVID) and Metascape^48^ with all GO terms considered significant at *p* < 0.05.

### MicroRNA analysis in MDE

MDE were analyzed for the expression of 13 miRNAs (Let-7e-5p, miR100-5p, miR-101-3p, miR-125a-5p, miR-125b-5p, miR-27a-3p, miR-27b-3p, miR-29a-3p, miR-34a-5p, miR-378a-3p, miR-423-5p, and miR-708-5p) related to mitochondrial function, with real-time PCR using TaqMan assays. cel-miR-39-3p was used as an external normalization control. Isolation of total RNA, including miRNA, and cDNA synthesis, was performed with MDE using our published protocol^7,13,46^. Prepared cDNA was diluted 3-fold, and 1µl of cDNA was used for the qPCR analysis using miRNAs specific TaqMan Advanced miRNA Assay (20X) in the final 10µl reaction. An equal amount of total RNA was used for each sample for cDNA synthesis, and an equal volume of cDNA was used for qPCR expression analysis. Relative expression of different miRNAs in each sample was normalized with external control (cel-miR-39-3p) to calculate ΔCt values.

Further, to analyze the MitoFunction score, the median value of each miRNA of all participants were determined and subtracted from the ΔCt values (normalized by cel-miR-39) to obtain the deviation from the median. The change in ΔCt value for each miRNA from the median (in any direction; increase or decrease) was scored as follows: 0-1= score 0, 1-2= score 1, 2-3= score 2, more than 3= score 3. The cumulative score from all miRNAs was presented as Mito-Function score.

### Enzyme-Linked Immunosorbent Assay (ELISA)

The level of peroxisome proliferator-activated receptor gamma (PPAR-γ) in MDE was analyzed using ELISA (MyBiosource, San Diego, CA, USA) after their lysis using 10X RIPA buffer following the manufacturer’s recommendation. The protein level in MDE lysate was measured by bicinchoninic acid assay (BCA) method. The concentration of PPAR-γ was calculated from the standards after normalization with the protein concentration.

### Statistical Analysis

Population characteristics were first compared between the two groups assigned to the laboratories at the UPitt and WFSM, and then by resilience status, using independent t-test and chi square when appropriate. EV characterization was analyzed by resilient group with a two-tailed unpaired t-test (GraphPad Prism v10.0.3). Biological function terms were reduced and visualized utilizing REVIGO, reduce+visualize Gene Ontology, (v 1.8.1). Comparisons of molecular cargo across groups were first unadjusted and then adjusted for demographics using linear regression models (SPSS 31.0).

## Results

### Health ABC participant characterization

Among the participants included in this analysis, 53% were women and 35% were Black, with an average age of 74 years. Mobility resilient was defined as at least one locomotor risk factor (pain, high BMI, low forced expiratory volume, peripheral artery disease) and a gait speed >1.0m/sec, while non-resilient individuals were defined as one locomotor risk factor and gait speed <1.0m/sec. Participants used for total serum circulating EVs and MDE did not significantly differ on any of the demographic characteristics examined (**Table 1**), consistent with the randomized assignment and supporting the comparisons of results between studies. As expected, based on the design of the study, resilient status was related to faster gait speed, and this was significant in both studies (**Table 2**). Additionally, and in both studies, resilient was associated with higher performance in the two cognitive tests (DSST, 3MS), and muscle strength (**Table 2**); these characteristics have been related with gait speed in prior studies^49,50^. Additionally, there was a statistically significant association between resilient status and sex in the MDE study (resilient were more likely to be women, **Table 2**) and by race in small ncRNA of total circulating EVs (resilient were more likely to be white, **Table 2**), thus these variables were considered as covariates in the analyses of EV markers. We started by isolating total circulating serum EVs from Health ABC participants classified as mobility resilient and non-resilient.

**Table 1.** Participants Characteristics for the experiments conducted at Site 1-Wake Forest University and Site 2-University of Pittsburgh.

| | Mean ±SD or n (%) | | p value |
| --- | --- | --- | --- |
|  | Site 1<br>N=21 | Site 2<br>N=24 |  |
| Age, years | 73.05±2.60 | 74.13±3.43 | 0.24 |
| Sex, women | 11 (52) | 13 (54) | 0.91 |
| Race, Non-Hispanic White | 14 (67) | 14 (58) | 0.76 |
| Gait Speed, m/sec | 1.30±0.56 | 1.07±0.27 | 0.11 |
| Comorbidities, number | 1.81±0.75 | 2.05±1.09 | 0.41 |
| Time spent walking (min/week) | 178.81±276.64 | 94.62±149.68 | 0.23 |
| Digit Symbol Substitution Test, points | 34.38± 12.53 | 35.25±19.84 | 0.86 |
| Modified Mini Mental Status Exam, points | 90.43±5.91 | 86.92±10.40 | 0.17 |
| Muscle Strength, nm | 92.34±34.02 | 82.94±34.73 | 0.42 |

**Table 2.**
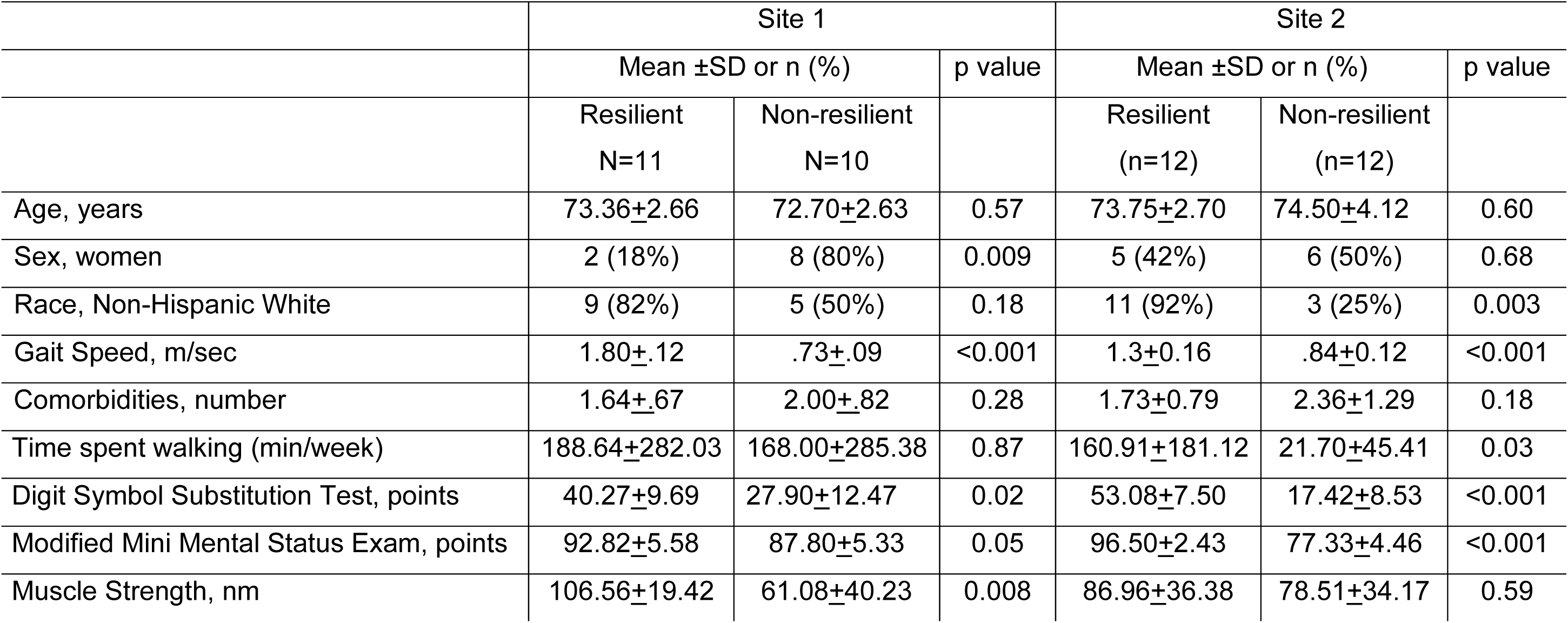
Participants Characteristics in Resilient and Non-resilient, for Site 1-Wake Forest University and Site 2-University of Pittsburgh.

|  | Site 1 |  |  | Site 2 |  |  |
| --- | --- | --- | --- | --- | --- | --- |
| | Mean ±SD or n (%) | | p value | Mean ±SD or n (%) | | p value |
|  | Resilient<br>N=11 | Non-resilient<br>N=10 |  | Resilient<br>(n=12) | Non-resilient<br>(n=12) |  |
| Age, years | 73.36±2.66 | 72.70±2.63 | 0.57 | 73.75±2.70 | 74.50±4.12 | 0.60 |
| Sex, women | 2 (18%) | 8 (80%) | 0.009 | 5 (42%) | 6 (50%) | 0.68 |
| Race, Non-Hispanic White | 9 (82%) | 5 (50%) | 0.18 | 11 (92%) | 3 (25%) | 0.003 |
| Gait Speed, m/sec | 1.80±.12 | .73±.09 | <0.001 | 1.3±0.16 | .84±0.12 | <0.001 |
| Comorbidities, number | 1.64±.67 | 2.00±.82 | 0.28 | 1.73±0.79 | 2.36±1.29 | 0.18 |
| Time spent walking (min/week) | 188.64±282.03 | 168.00±285.38 | 0.87 | 160.91±181.12 | 21.70±45.41 | 0.03 |
| Digit Symbol Substitution Test, points | 40.27±9.69 | 27.90±12.47 | 0.02 | 53.08±7.50 | 17.42±8.53 | <0.001 |
| Modified Mini Mental Status Exam, points | 92.82±5.58 | 87.80±5.33 | 0.05 | 96.50±2.43 | 77.33±4.46 | <0.001 |
| Muscle Strength, nm | 106.56±19.42 | 61.08±40.23 | 0.008 | 86.96±36.38 | 78.51±34.17 | 0.59 |

### Characterization of total circulating serum EV and small ncRNA cargos

We first characterized the isolated serum EVs based on size, concentration, protein content, and RNA concentration. Nanoparticle tracking analysis (NTA) showed enrichment of serum EVs with similar distribution of size in both the mobility resilient and non-resilient participants (**Fig. 1A**). The NTA analysis showed no significant difference in the average size of isolated serum EV between the groups, with the predominance of particles measured between 50 nm and 200 nm which indicates enrichment of small EVs (**Fig. 1B**). Similarly, there was no significant difference in the average EVs concentration between groups, with the resilient group having 5.1×10^9^ particles/ml and non-resilient group having 5.7×10^9^ particles/ml (**Fig. 1C**). The enrichment of small EVs from serum samples was confirmed by Western blotting using canonical EVs markers CD63 and CD81 (**Fig. 1D**). Furthermore, transmission electron microscopy (TEM) further confirmed the presence of intact EVs isolated from the serum, with enrichment of small EVs (**Fig. 1E**). These findings demonstrate successful isolation and characterization of serum EVs from these participants, with no significant difference in their biophysical properties.

**Figure 1.**
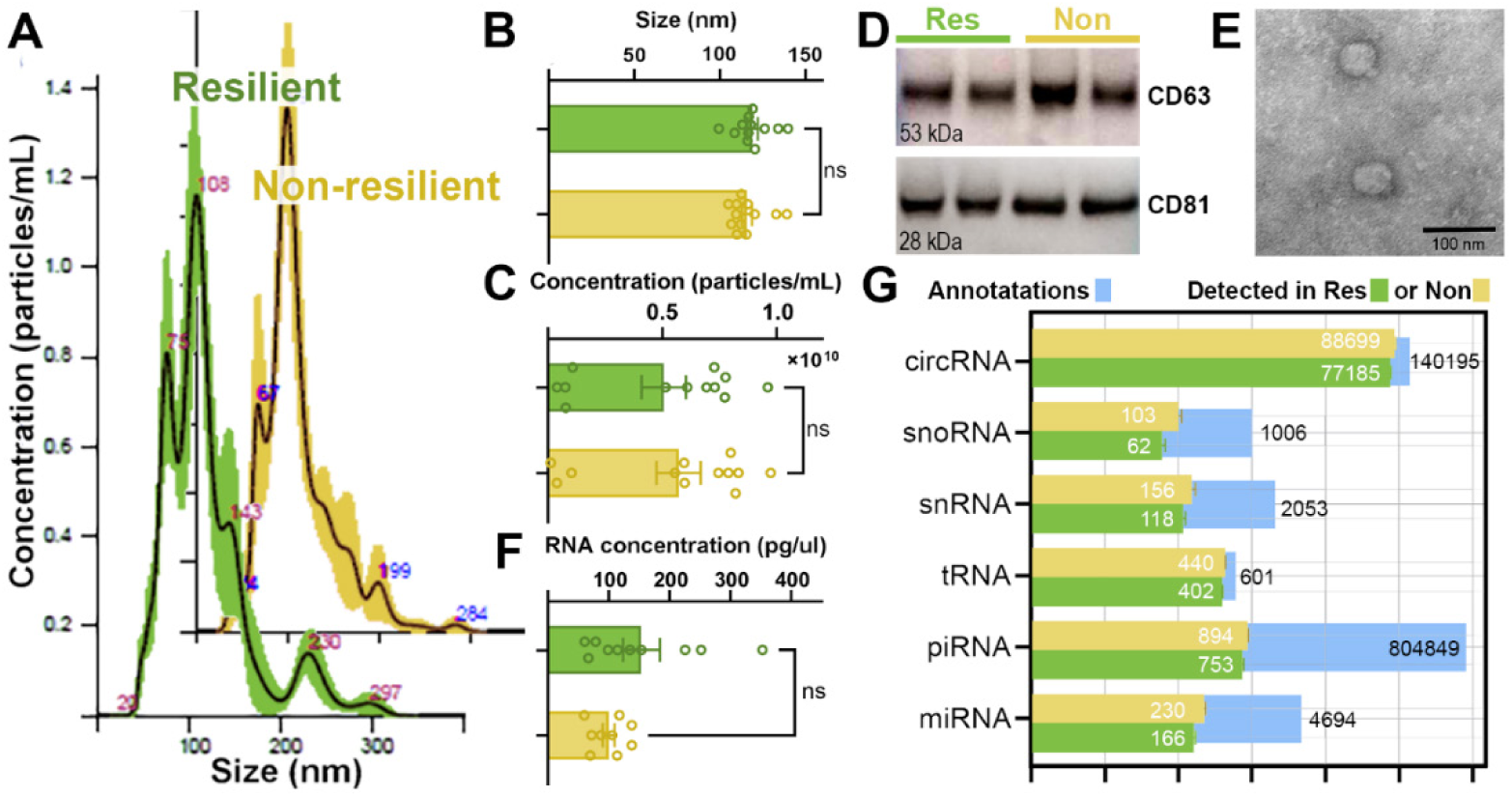
Characterization and quantification of total circulating EVs isolated from the serum of mobility Resilient and Non-resilient individuals. **(A)** Isolated total circulating serum EV particle size distribution profiles were measured by NTA from resilient (Res, green) and non-resilient (Non, yellow) individuals showing similar size distribution in the study participants. Bar graphs representing EV size **(B)** and concentration **(C)**, as measured by NTA analysis for serum EV isolated from resilient and non-resilient groups showing no significant difference between groups. **(D)** Representative western blot images of classical EV markers CD63 and CD81 showing no difference in thier levels between serum EVs isolated from the cohort. **(E)** Representative TEM image to visualize size and morphology of EVs isolated from participants serum. **(F)** Bar graph showing no significant difference in the concentration of RNA isolated from the serum EVs. (G) Bar plot showing the number of small ncRNA transcripts annotated from serum EVs of the resilient and non-resilient groups. Blue bars represent the total number of possible circRNA, snoRNA, snRNA, tRNA, piRNA and miRNA annotations in COMPSRA. Note that there was a high percentage of annotated small ncRNAs and no significant difference in the numbers of annotated small ncRNAs when comparing the two groups. Bar graphs represent mean ± SEM; n = 12 serum samples/group. Statistical significance was determined by unpaired t-test. ns, not significant.

Following serum EVs characterization, total RNA was isolated from the EVs, and there was no significant difference in the amounts of total RNA between the resilient and non-resilient groups (**Fig. 1F**). Most of the isolated RNA displayed a size between 50 to 200 nucleotides which is a characteristic of small ncRNA. The sequencing data were analyzed for the abundance of six distinct small ncRNA classes: circRNA, snoRNA, snRNA, miRNA, piRNA and tRNA annotated with COMPSRA^51^. We observed no differences between the groups in the number of annotated small ncRNAs identified in the serum EVs (**Fig. 1G**). These findings support that we efficiently isolated total circulating serum EVs and sequenced and annotated the small ncRNA cargos. There was no significant difference in the types or number of EVs or small ncRNA species annotated between the mobility resilient and non-resilient samples.

### Differential analysis of enriched small ncRNAs from total circulating serum EV of mobility resilient individuals

Subsequent analysis on the differential abundance of the small ncRNA cargos was performed with the negative binomial distribution (DESeq2) R package to determine statistical enrichment of these cargos in the total circulating serum EVs comparing resilient and non-resilient individuals. Among the six classes of small ncRNAs, we found several species which were differentially abundant in the serum EVs between resilient and non-resilient groups (**Fig. 2A**). The circRNAs had the highest number of species which showed differential enrichment, with 720 increased in non-resilient and 1588 increased in serum EVs from resilient participants (**Fig. 2A**). In resilient individuals, there was a higher percentage of species significantly enriched belonging to tRNAs (11.2%), snRNAs (5.1%), and miRNAs (5.1%) while non-resilient individuals, a high percentage was changed in snoRNA (19.5%), miRNAs (8.6%) and tRNAs (6.6%) (**Fig. 2B**). We observed that two tRNAs: Thr-CGT-2-1 and Tyr-GTA-9-1 were enriched in resilient group. Of particular interest, several SNORD116 family members were enriched in EVs of the non-resilient group (**Fig. 2A, snoRNA**). These SNORD116 species are highly expressed in neurons, and we have shown that they are increased in plasma EVs of Alzheimer’s disease (AD) patients compared to healthy controls^4^. Similarly, we observed an increased abundance of piR_25781, piR_28187 and piR_28188 in EVs of the non-resilient cohort (**Fig. 2B**), which showed similar increased levels in the brains of AD patients^52,53^. To reduce the dimensionality of the data and visualize differences in the types and abundance of annotated small ncRNA cargos from these two cohort EV samples, we performed a Principal Component Analysis (PCA) analysis. PCA plot of the small ncRNA abundance levels for the resilient and non-resilient segregated into two separate groups, forming distinct clusters. This finding indicates unique abundance profiles between the cohorts (**Fig. 2C**). Many of the significantly differential abundant miRNAs (mir-370-3p, 204-5p, 181b-5p, and 127-3p) in serum EVs between the two groups were previously shown to have enriched expression in brain tissue (**Fig. 2D**), as identified in published small ncRNA tissue atlases^54–56^. Furthermore, these miRNAs have key target genes associated with health brain aging. Interestingly, mir1253 which is enriched in the serum EVs of the resilient group showed skeletal muscle tissue specific expression in the same published atlases. This suggests that serum EVs as a source for assessing changes in small ncRNA related pathways in both brain and peripheral tissue.

**Figure 2.**
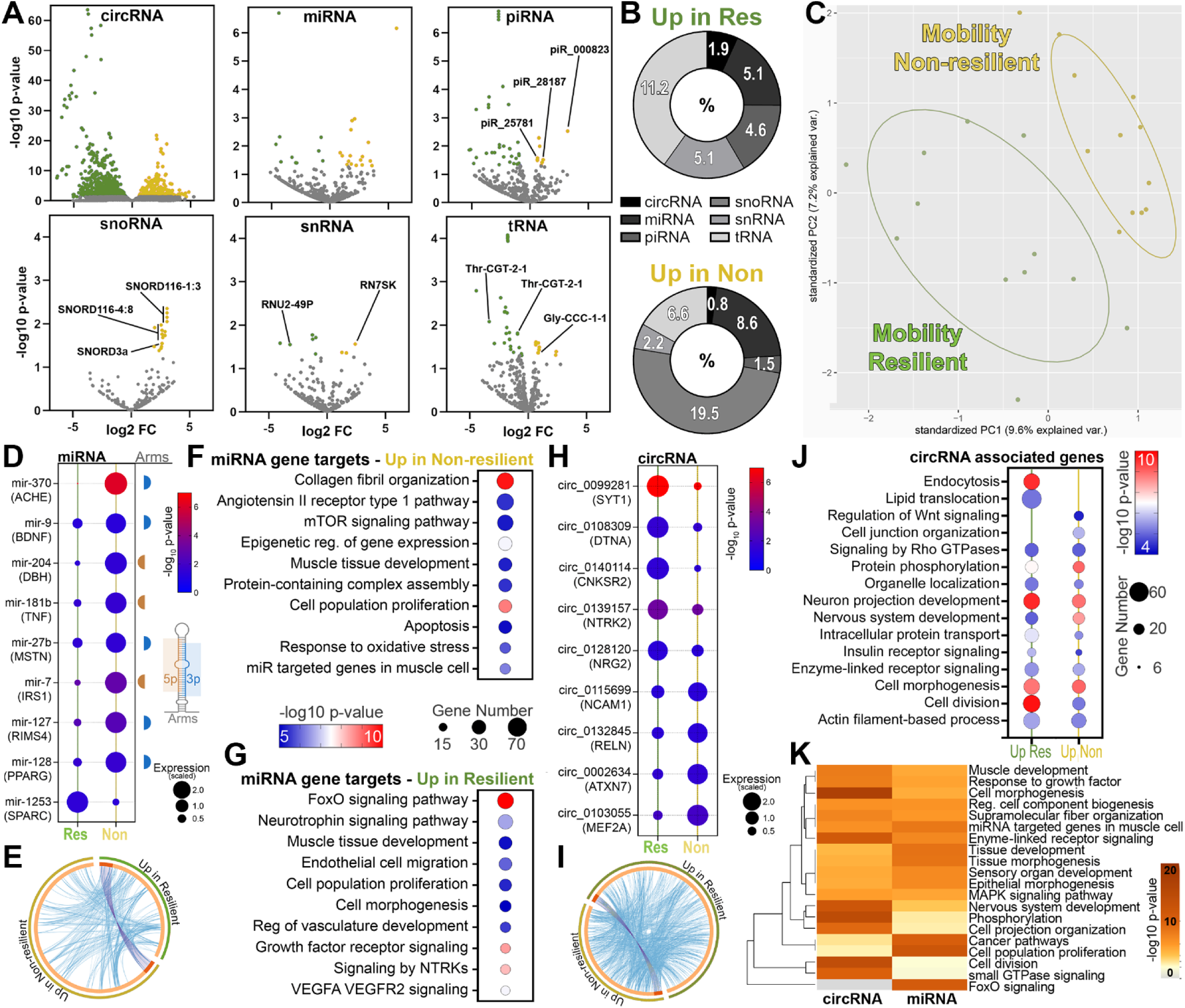
Unique small ncRNA profile of total circulating serum EVs associated with mobility status. **(A)** Volcano plots showing differential enrichment of circRNAs, miRNAs, piRNAs, snoRNAs, snRNAs, and tRNAs from total circulating serum EVs of resilient (Res) compared to non-resilient (Non) individuals. **(B)** Donut plot showing distribution of significantly affected small ncRNA species compared to total annotated in either resilient or non-resilient group. **(C)** PCA plot showing unique sncRNA profiles for the two experimental groups. **(D)** Bubble plot showing abundance of selected significantly affected miRNAs, which show brain tissue enrichment, except miRNA-1253, which is muscle tissue enriched. Select target gene below the species and identified arm to the right of the plot. The color of the dot represents the significance and size of the dot represents relative expression. **(E)** Circos plot shows overlap of Targetscan defined target genes associated with miRNAs that were enriched in the serum EVs of resilient or non-resilient individuals. Dark orange color represents the genes that are shared while light orange shows unique genes in the two groups. Purple lines link the shared genes, and blue lines link unique genes that belong to the same ontology term. **(F-G)** Bubble plots show the biological terms associated with the gene targets of enriched miRNAs in serum of EVs of non-resilient **(F)** and resilient **(G)** participants. The color of dot represents the significance and size of dot represents the number of genes in the ontology term. **(H)** Bubble plot showing abundance of selected significantly altered circRNAs which show brain tissue enrichment. Selected associated gene below the species **(I)** Circos plot shows overlap of circBase defined circRNA associated genes that were enriched in the serum EVs of resilient or non-resilient individuals. Dark orange color represents the genes that are shared while light orange shows unique genes in the two groups. Purple lines link the shared genes, and blue lines link unique genes that belong to the same ontology term. **(J)** Bubble plots show the biological terms of the associated gene of enriched circRNAs in the serum of EVs of resilient and non-resilient participants. **(K)** Dendrogram shows selected biological terms with the best p-value that were common when assessing circRNA and miRNA associated input genes. The heatmap cells are colored by their p-values. n = 12 individuals /group.

To better understand the biological significance in these enrichment patterns in the serum EVs, we identified target genes of the differentially enriched miRNAs with TargetScan Human. Using the 27 significantly affected miRNAs (10 increased in resilient group and 17 increased in non-resilient group), we determined 249 target genes associated with the resilient group and 471 associated with the non-resilient group, with 24 target genes that were common targets of significantly enriched miRNAs from both cohorts (**Fig. 2E, purple lines**). Furthermore, there was a low concurrence of functional overlaps among the target genes associated with the two cohorts, suggesting changes in unique biological functions between the two groups associated with changes in the miRNA cargo levels (**Fig. 2E, blue lines**). We then submitted the miRNA target gene lists associated with the resilient and non-resilient groups to Metascape aiming to uncover the potential functional roles of these miRNAs and their gene targets. In the non-resilient group, we observed biological functions of the target genes associated with Collagen fibril organization, mTOR signaling pathways, response to oxidative stress, cell proliferation and apoptosis, muscle tissue development, and miRNA targeted genes in muscle cells (**Fig. 2F**). In resilient individuals, target genes were associated with FoxO signaling, neurotrophin and growth factor signaling, muscle and vascular development, and endothelial cell migration (**Fig. 2G**). We also found several circRNA that had differential abundance between the two groups with enriched expression in the brain (**Fig. 2H**)^57,58^. We identified 304 associated genes from the enriched circRNAs of the non-resilient group and 551 from the resilient group. There were 19 associated genes that were in both lists, and the genes associated with the two groups showed a higher level of functional overlap compared to miRNA target genes suggesting a more shared biological function between the circRNA cargos of the two cohorts (**Fig. 2I**). The resilient group had unique biological functions from the circRNA associated genes linked to endocytosis and lipid transport while the non-resilient group had unique functions linked to cell junction and Rho GTPase signaling. The circRNA associated genes from both groups shared a lot of biological functions such as organelle organization, nervous system development, insulin and enzyme linked signaling, and cell morphogenesis and division (**Fig. 2I**). Comparative analysis of the miRNA and circRNA associated genes highlights the potential role of these cargos in biological functions and pathways that may contribute to the phenotypes of the resilient and non-resilient individuals (**Fig. 2K**). The hierarchical clustering of statistically enriched terms demonstrates that these serum EV cargos have shared roles in biological functions such as muscle and nervous system development, response to growth factors, cell projections, proliferation and morphogenesis, response to growth factors and cell component biogenesis. Overall, analysis of the small ncRNA cargo from serum EVs of resilient compared to non-resilient group suggest packaging of these cargos is not random and may be involved in aspects of phenotypes, disease onset, and progression. These results also show a small ncRNA mobility-related signature of serum EVs of non-resilient individuals, which this analysis suggests captures changes in disease mechanisms related to brain and muscle functions.

### MDE display unique molecular signatures of mitochondrial function according to mobility status

To better elucidate the ability of EVs cargo to reveal skeletal muscle ergogenic states, we isolated total circulating serum EVs from the serum of 21 Health ABC participants, including mobility resilient and non-resilient adults, followed by MDE isolation, as mentioned in methods^13,59^. The Nano-flow cytometry analysis showed no significant difference in surface expression of mitochondrial markers TOM20, VDAC, mtCox-2, and PHD (**Fig. 3A**) in total circulating serum EVs between the two groups. Importantly, for MDE, the resilient group displayed higher surface expression of mitochondrial markers, including TOM20 and VDAC, compared to the non-resilient group (**Fig. 3B**). However, we did not observe any significant difference in the surface expression of mtCox-2 and PDH between the two groups in both total circulating serum EVs and MDE. Next, we characterized the cargos of MDE, across groups for miRNA expression and MitoFunction Score. First, we analyzed the expression of a panel of 13 miRNAs, regulating mitochondrial function to determine their association with mobility resilience, in MDE. Second, the MitoFunction Score was calculated, based upon the change in the expression of these miRNAs (**Supplementary Fig. 1**), which displayed significantly higher score for non-resilient older adults as compared to resilient older adults (**Fig. 4**). Lastly, we characterized the expression of PPAR-γ, a nuclear receptor, which when activated promotes mitochondrial biogenesis^60^. As shown in Fig. 4B, we observed higher levels of PPAR-γ in the MDE of resilient group.

**Figure 3.**
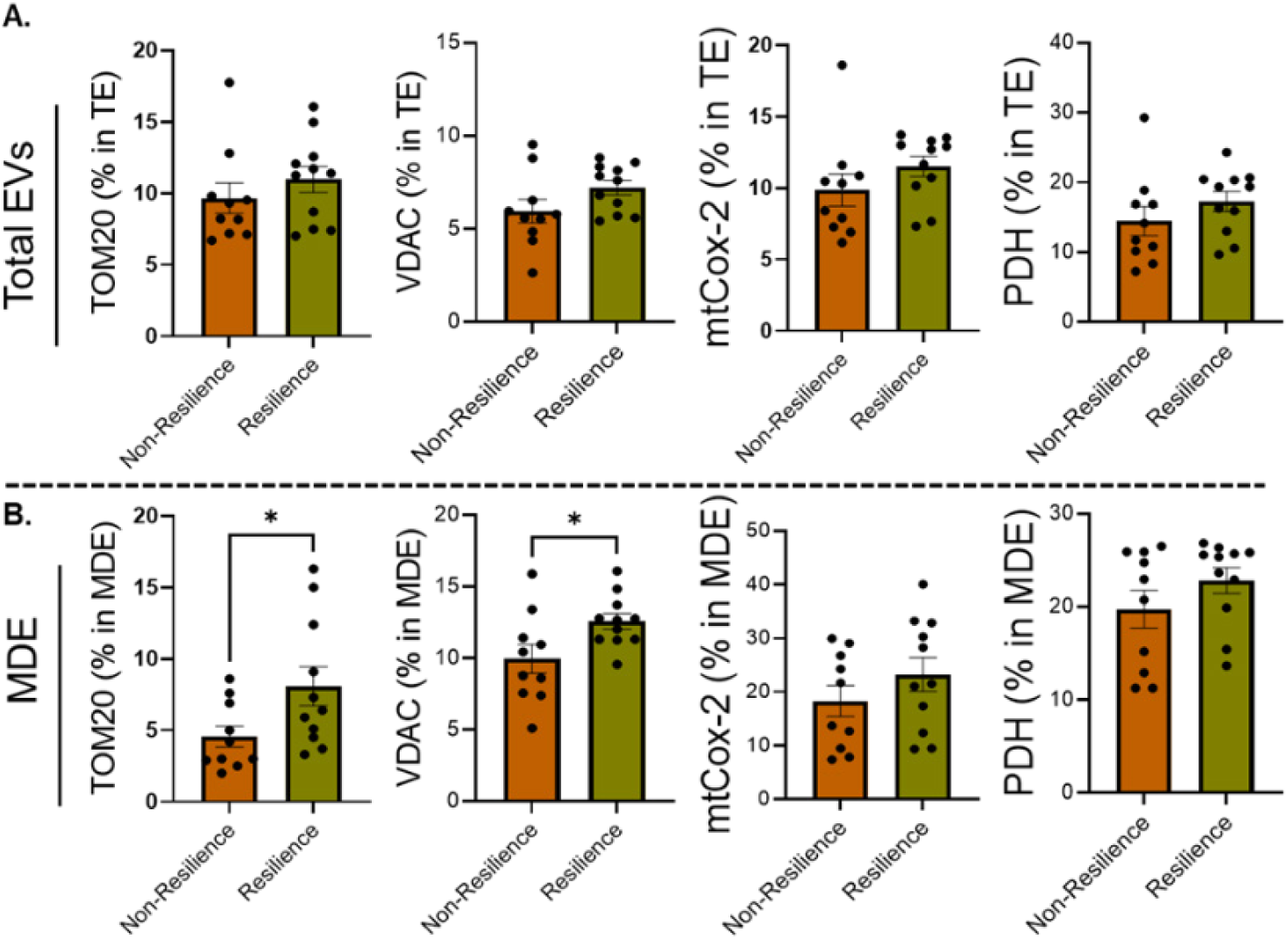
Assessment of mitochondrial markers in total serum EVs and muscle-derived EVs (MDE). Mitochondrial markers on the EVs surface was measured using nanoflow cytometry. **(A)** Serum samples, from mobility non-resilient (n=10) and resilient (n=11) participants were processed to isolate total EVs and analyzed for the surface expression of TOM20, VDAC, mtCox-2, and PDH on their surface. **(B)** MDE, isolated from total EVs, were assessed for the surface expression of same mitochondrial markers (TOM20, VDAC, mtCox-2, and PDH). Statistical significance was determined by unpaired t-test. *, p<005. TOM20: translocase of outer mitochondrial membrane 20; VDAC: voltage-dependent anion channel; mtCox-2 mitochondrial Cytochrome c oxidase subunit II; PDH: Pyruvate dehydrogenase.

**Figure 4.**
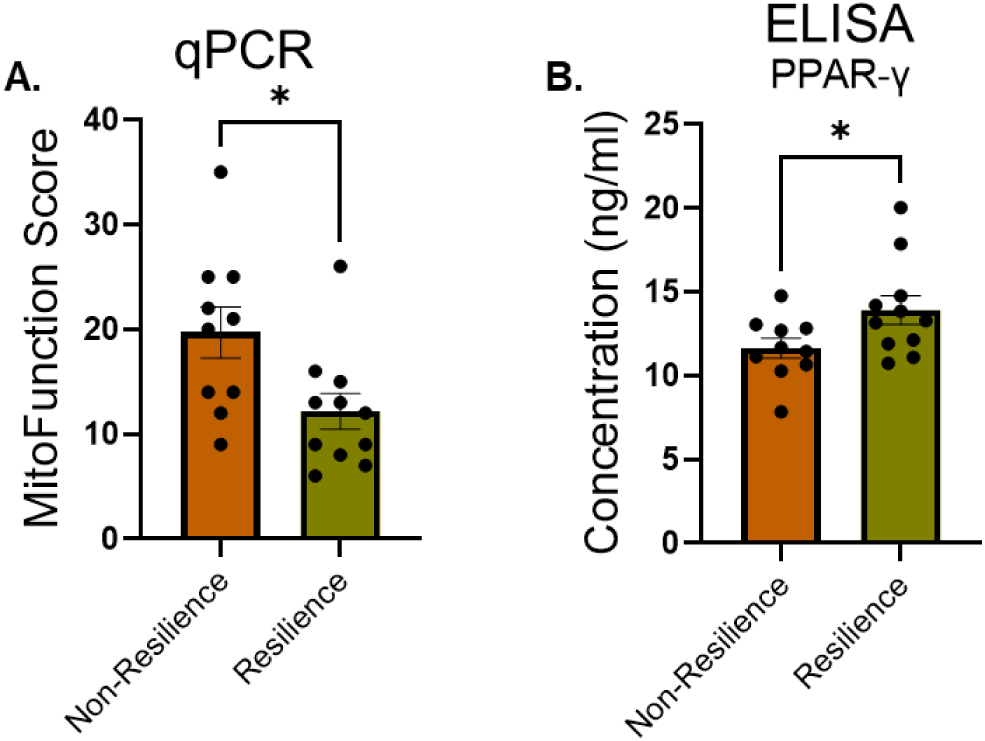
Assessment of MDE cargos for biomarkers of mitochondiral function. **(A)** MDE were analyzed for 13 miRNAs by RT-PCR and based on their expression levels the MitoFunction Score was calculated for non-resilient and resiliene groups as described in the methods. **(B)** The levels of PPAR-γ was analyzed using ELISA in MDE from non-resilient and resilient groups. Statistical significance was determined by unpaired t-test.*, p<005

We correlated levels of the assessed MDE cargos with clinical phenotypes. Age and gender adjusted correlations between these MDE cargos and gait speed were statistically significant for TOM20, miR34a-5p and miR27b-3p (adjusted correlation coefficients [p values]: 0.50 [p=0.028], - 0.67 [p=0.002], -0.50 [p=0.03]), but not PPAR-γ Additionally, TOM20 and miR34a-5p were also associated with muscle strength, (adjusted correlation coefficients [p values]: 0.58 [p=0.03], - 0.73 [p=0.003]). TOM20 was positively correlated with modified mini-mental exam (3MS) score (adjusted correlation coefficients [p values]: 0.49 [p=0.033]) and miR34a was negatively correlated with Digit Symbol Substitution Test (DSST) score (adjusted correlation coefficients [p values]: -0.57 [p=0.014]).

## Discussion

Our study is the first to provide a comprehensive molecular characterization of total serum EVs and MDE in older adults with differing mobility resilience. By integrating small ncRNA profiling and mitochondrial marker analysis, we demonstrate that EVs carry distinct systemic and muscle-specific signatures reflective of biological aging and functional capacity. The identification of differentially enriched miRNAs, circRNAs, and mitochondrial proteins in EVs from resilient versus non-resilient individuals highlights their potential as non-invasive biomarkers for mobility outcomes. We found that small ncRNA cargos were differentially abundant in older adults who were resilient compared to non-resilient, with the highest differences in tRNA, miRNA, and snoRNA members. The associated genes of differentially abundant miRNAs and circRNAs in the two cohorts indicated variations in biological processes related to muscle and nervous system development, metabolic signaling, and cellular responses to stress. Focused analysis of MDE exhibited enhanced mitochondrial markers in resilient individuals, suggesting a role in preserving muscle bioenergetics. Our study demonstrates the possibility of small ncRNA and protein EVs cargos as biomarkers for mobility resilience in older adults and that these potential biomarkers are associated with biological functions important in muscle and brain health. This work advances the field of precision geroscience by linking EV mediated molecular communication to functional aging phenotypes and lays the groundwork for future biomarker-driven strategies to promote mobility resilience and healthy aging.

Our initial characterization of total circulating serum EVs revealed no significant differences in size, concentration, or canonical EV markers (CD63, CD81) between resilient versus non-resilient group, suggesting that the observed molecular differences are not due to EVs quantity or quality, but rather to selective cargo loading. Further, small ncRNA profiling identified a diverse repertoire of RNA species, consistent with recent reports highlighting their enrichment in circulating EVs and roles in aging and cellular stress responses^61–63^. Among the miRNAs in total circulating serum EVs in our data, 17 were enriched in the resilient group and 10 enriched in the non-resilient group. Interestingly, in the resilient group, 8 of 17 significantly differential abundant miRNAs show higher expression in the brain compared to other tissues according to small ncRNA tissue atlases^54–56^. This contrasts with the non-resilient group, which had no increased miRNAs that show brain enriched expression. Furthermore, miR-1253, which is highly expressed in muscle tissue and has been shown to impair proliferation and induce apoptosis^64^, was significantly increased in the total circulating serum EVs of non-resilient individuals. This highlights the importance of miRNA EVs cargos in the cross talk of brain and peripheral tissues and could provide a mechanism for mobility resilience with aging.

Impaired metabolic flexibility of skeletal muscle likely contributes to the development of certain chronic diseases, including obesity and type 2 diabetes, and affects mobility resilience with age^65^. In our data, miR-183, a master regulator of metabolic homeostasis in skeletal muscle, was enriched with the highest fold change in serum EVs isolated from mobility non-resilient individuals. Increased miR-183 level could decrease β-oxidation and lipolysis in skeletal muscle and facilitate glucose utilization by impairing phosphorylation of PDHA1 via targeting FoxO1^66^. Furthermore, previous studies have shown that decreasing miR-183 enhances fat catabolism in skeletal muscle and has a beneficial effect for muscle health after a high fat diet challenge^66^. It is generally accepted that decreased β-oxidation and higher utilization of glucose stores in skeletal muscle is associated with metabolic diseases^67,68^, which, along with changes in lipid metabolism, could contribute to mobility decline in the non-resilient individuals.

Interestingly, we observed significant increases in miR-3168 and miR-126 in EVs of non-resilient individuals, which may impact vascular health contributing to mobility phenotype. Overexpression of miR-3168 and miR-126 impairs angiogenesis by targeting *BMPR2*^69^ and *HIF-1α*^70^, respectively. Crosstalk between skeletal muscle and vascular endothelial cells is essential for the coordinated delivery of oxygen and nutrients to active tissues. This interaction, when coupled with angiogenic signaling, enhances capillary density and perfusion, thereby supporting improved exercise endurance and metabolic efficiency. This link between skeletal metabolic functions and miRNA EV cargos is further exemplified by significant increases in miR-29a-3p in total circulating serum EVs of resilient individuals. The level of miR-29a-3p in total circulating serum EVs is reported to be increased after resistance training and mediates changes in energy metabolism in muscle^71^. We also observed increased levels of miR-29a-3p in MDE (though not statistically significant) from resilient individuals, which contributes to the diminished MitoFunction Score of non-resilient individuals. There is strong evidence that mitochondrial dysfunction leads to alteration in skeletal muscle metabolism, a hallmark of aging and a root cause of progressive loss of skeletal muscle function and CNS dysfunction, both of which contribute to mobility decline and ultimately physical disability^72^.

In total circulating serum EVs, the abundance of tRNAs was also significantly impacted by mobility resilience in older individuals. It remains largely unknown how physiological and pathological conditions regulate tRNAs and their derivatives, which are important components of the protein synthesis machinery in muscle. Differential expression analysis revealed significant alterations in ncRNA abundance between resilient and non-resilient groups, with circRNAs showing the greatest number of differentially enriched species. Of particular interest, two tRNAs, Thr-CGT-2-1 and Tyr-GTA-9-1, were enriched in resilient individuals. These tRNAs have previously been associated with better muscle strength and physical performance in older adults^63^, suggesting their potential role in promoting mobility resilience. Interpretation of these findings could indirectly be linked to anabolic effects and muscle protein synthesis given the critical role of leucine (Leu). Notably, Leucyl-tRNA synthetase is an intracellular sensor that transfers Leu to cognate tRNAs to form tRNA-Leu species and this activates mTOR signaling, serving to activate the protein synthesis and anabolic processes^73^. There are no studies that have directly investigated tRNA-Leu species and muscle-related metabolism, and therefore, the underlying mechanisms remain unclear. Our study is in agreement with a similar result that circulating tRNA-Leu species are significantly affected in older individuals with low muscle strength and physical performance^63^, while, in our study, we demonstrate that the source of these circulating tRNAs are the serum EVs.

Mobility decline with age is the result of the interplay of progressive loss of skeletal muscle functions, CNS dysfunction, and altered brain-muscle communication. We detected several snoRNAs and piRNAs cargos that were significantly enriched in total circulating serum EVs from the non-resilient group, that have been previously linked to neurodegeneration. Several members of the SNOD116 family were increased in non-resilient individuals. SNORD116 is predominantly expressed in the brain, and we previously demonstrated increased levels of members of the SNORD116 family in the plasma EVs of AD patients^4^. SNORD116 functions by regulating the stability of mRNA targets, such as Nhlh2, a transcription factor involved in energy homeostasis and metabolic function^74^. Similarly, we observed increased levels of piR_25781, piR_28187 and piR_28188 in the total circulating serum EVs of non-resilient individuals. These piRNAs are increased in brain tissue of AD patients^52,53^. These results highlight that biomarkers associated with neurodegeneration are also increased in the total circulating serum EVs of non-resilient individuals. This suggests the CNS may be the origin for these EVs and that alterations in brain EV cargos could contribute to changes in mobility through brain-muscle cross talk. This also highlights a limitation of utilizing total circulating serum EVs as a liquid biopsy. Total circulating serum EVs contain EVs released by multiple organs, including the brain, where there are mechanisms allowing for the transport of EVs across brain barriers. The non-resilient compared to resilient individuals in our study exhibited significantly worsened cognitive testing scores as assessed by modified mini-mental exam (3MS) and digit symbol substitution test (DSST) which could not only contribute to the unique small ncRNA signature observed from the total circulation serum EVs but also contribute to mobility decline with age. More research is needed to better define markers of brain released EVs found in peripheral biopsies that may contribute to brain-muscle communication and associated with the pathology of mobility decline with age.

Skeletal muscle is one of the largest organs in the body and can release EVs into the circulating serum, which can play a significant role in communication of multiple systems. In this study, we demonstrate that EVs derived from muscle contain not only differential abundant miRNAs that are important in mitochondrial function but also display changes in protein cargoes that are important in muscle metabolic functions. In MDE, resilient individuals showed significantly higher expression of mitochondrial markers, indicating enhanced mitochondrial content and potential bioenergetic capacity. This finding aligns with the concept that mitochondrial health is a key determinant of muscle function and resilience in aging^75–77^. MDE could serve as a vehicle for mitochondrial proteins in intercellular communication, delivering metabolic signals that support tissue homeostasis and adaptation to stress. Importantly, the levels of both miR27b-3p and miR34a-5p in MDE showed a strong negative correlation to mobility phenotypes (gait speed or muscle strength), suggesting a direct link between MDE cargos and mobility resilence. Further analysis of the MitoFunction Score demonstrated an increase in non-resilience individuals. Since the MitoFunction score is not based on the directionality of change in expression of its contributing miRNAs, its increase in non-resilience individuals may reflect a compensatory response to mitochondrial dysfunction, as mitochondrial-related miRNAs are known to play important regulatory role in conditions of oxidative stress and impaired bioenergetics^78^. Additionally, higher levels of PPAR-γ in MDE from resilient individuals suggest a state of relatively higher mitochondrial biogenesis. Interestingly, we also observed that two of the miRNAs which were enriched in total circulating serum EVs from non-resilience individuals, have target genes that are associated with *VDAC* (miR-183-3p) and *PPARG* (miR-27b-3p), suggesting that in individuals with declining mobility not only could these protein cargos be diminished in EVs but expression of their associated genes could be repressed by these miRNA cargos in target tissues. Importantly, we observed correlations in the levels of TOM20 in MDE with mobility phenotypes in this cohort. These correlations between protein cargo and previously reported associations with miRNA cargo underscore the potential of MDE as biomarkers for mobility decline. Interestingly, we also observed correlations between TOM20 and miR-34a-5p and cognitive assessments in this cohort, which supports the idea of EVs of muscle origin can modulate cognitive functions in older adults. To validate and expand upon these findings, further investigation of MDE cargo should be conducted in a larger, well-characterized cohort. These findings also support the hypothesis that MDE carry molecular signatures indicative of skeletal muscle ergogenic states and that these signatures differ according to mobility resilience. The enhanced mitochondrial marker expression in resilient individuals may reflect preserved mitochondrial integrity and adaptive capacity, contributing to better physical performance.

The results of this study demonstrate that total circulating serum EVs from resilient older adults have a unique small ncRNA signature related to genes important in brain and muscle development, metabolic homeostasis, and cellular response to stress. Further MDE revealed differential expression of TOM20, VDAC and PPAR-γ as well as MitoFunction score between the two groups. In addition, TOM20, miR-34a-5p, miR-27b-3p isolated from MDE showed strong correlation to mobility phenotypes. Although the outcomes of much larger independent validation studies are difficult to predict, the results presented here point to a newly discovered association of EVs cargos with mobility resilience in aging individuals. This preliminary study supports a large cohort study examining the potential of total circulating serum EVs or EVs from brain and muscle origin as a novel source in search of biomarkers with high discriminatory power to differentiate changes in mobility associated with age. Moreover, the EV cargos offer a valuable source for monitoring biological processes impacted by interventions or therapies to improve mobility resilience.

## Conclusion

Together, our data suggests that total circulating serum EVs and MDE provide complementary insights into the biological underpinnings of mobility resilience. The small ncRNAs from serum EVs reflect systemic and CNS-related pathways, while MDE reveal muscle-specific mitochondrial signatures. Integrating both the molecular profiles offers a powerful approach to identify biomarkers and therapeutic targets for promoting resilience in aging. Moreover, the identification of specific ncRNAs and mitochondrial markers associated with resilience open avenues for targeted interventions. For example, enhancing the expression of protective tRNAs or circRNAs, or modulating miRNA networks involved in FoxO and neurotrophin signaling, may improve muscle and vascular health. Similarly, strategies to boost mitochondrial content in MDE, such as exercise, nutritional supplementation, or pharmacological activation of PPARγ, could enhance muscle bioenergetics and delay mobility decline. Conclusively, this study demonstrates that total circulating serum EVs, and MDE, carry molecular signatures reflective of mobility resilience in older adults. The differential enrichment of small ncRNAs and mitochondrial markers between resilient and non-resilient group highlights the potential of EVs as biomarkers and modulators of aging-related functional outcomes. By elucidating the molecular cargo of EVs, we pave the way for novel diagnostic and therapeutic strategies to promote healthy aging and preserve mobility.

## Data Availability

All data produced in the present study are available upon reasonable request to the authors or online at https://www.ncbi.nlm.nih.gov/geo/

https://www.ncbi.nlm.nih.gov/geo/

## Availability of data and materials

Raw data single-cell RNA-seq was submitted to GEO (https://www.ncbi.nlm.nih.gov/geo/) and will be publicly available as of the date of publication. This paper does not report the original code. Any additional information required to reanalyze the data reported in this paper is available from the corresponding author upon request.

## Abbreviations

3MS: modified mini-mental exam
AD: Alzheimer’s disease
BBB: blood-brain barrier circRNAs circular RNAs
CNS: central nervous system
DSST: digit symbol substitution test
EV: extracellular vesicles
MDE: muscle-derived EVs
miRNAs: microRNAs
mtCox2: cytochrome c oxidase subunit 2
ncRNA: non-coding RNA
NTA: nanoparticle tracking analysis
PDH: pyruvate dehydrogenase
piRNAs: PIWI-interacting RNAs
PPAR-γ: peroxisome proliferator-activated receptor gamma
SGCA: α-sarcoglycan
SMS: skeletal muscle system
snRNAs: small nuclear RNAs
snoRNA: small nucleolar RNAs
TEM: transmission electron microscopy
TOM20: translocase of outer mitochondrial membrane 20
tRNAs: transfer RNAs
VDAC: voltage-dependent anion channel

## Authors’ contributions

Conceptualization, N.F.F., C.R., F.A., G.D.; Methodology, N.F.F., A.K., G.D. ; Investigation, N.F.F., A.K., Y.S.,

M.S., S.S.; Formal analysis, N.F.F., A.K., C.R.; Visualization, N.F.F., A.K., C.R.; Writing – Original draft, N.F.F.,

A.K., C.R., G.D.; Writing – Review & Editing, N.F.F., A.K., Y.S., M.S., S.S., R.K., I.L., A.S., F.A., C.R., G.D.;

Supervision, N.F.F., C.R. G.D.; Project administration, N.F.F., C.R. G.D.; Resources, N.F.F., C.R. G.D.; Funding acquisition N.F.F., C.R. G.D.

## Ethics declarations

Ethics approval and consent to participate

All studies were approved by the University of Pittsburgh Institutional Review Board.

Consent for publication Not applicable.

## References

1 Cummings, S. R., Studenski, S. & Ferrucci, L. A diagnosis of dismobility--giving mobility clinical visibility: a Mobility Working Group recommendation. JAMA 311, 2061–2062 (2014). 10.1001/jama.2014.3033

2 Studenski, S. et al. Gait speed and survival in older adults. JAMA 305, 50–58 (2011). 10.1001/jama.2010.1923

3 Rosso, A. L., Studenski, S. A., Longstreth, W. T., Jr., Brach, J. S., Boudreau, R. M. & Rosano, C. Contributors to Poor Mobility in Older Adults: Integrating White Matter Hyperintensities and Conditions Affecting Other Systems. J Gerontol A Biol Sci Med Sci 72, 1246–1251 (2017). 10.1093/gerona/glw224

4 Fitz, N. F., Wang, J., Kamboh, M. I., Koldamova, R. & Lefterov, I. Small nucleolar RNAs in plasma extracellular vesicles and their discriminatory power as diagnostic biomarkers of Alzheimer’s disease. Neurobiol Dis 159, 105481 (2021). 10.1016/j.nbd.2021.105481

5 Sahu, A. et al. Regulation of aged skeletal muscle regeneration by circulating extracellular vesicles. Nat Aging 1, 1148–1161 (2021). 10.1038/s43587-021-00143-2

6 Mishra, S. et al. Circulating small extracellular vesicles as blood-based biomarkers of muscle health in aging nonhuman primates. Geroscience 47, 3709–3723 (2025). 10.1007/s11357-024-01439-y

7 Kumar, A. et al. MicroRNA expression in extracellular vesicles as a novel blood-based biomarker for Alzheimer’s disease. Alzheimers Dement 19, 4952–4966 (2023). 10.1002/alz.13055

8 Kumar, A. et al. Small extracellular vesicles in plasma reveal molecular effects of modified Mediterranean-ketogenic diet in participants with mild cognitive impairment. Brain Commun 4, fcac262 (2022). 10.1093/braincomms/fcac262

9 Welsh, J. A. et al. Minimal information for studies of extracellular vesicles (MISEV2023): From basic to advanced approaches. J Extracell Vesicles 13, e12404 (2024). 10.1002/jev2.12404

10 Cleary, J. A., Kumar, A., Craft, S. & Deep, G. Neuron-derived extracellular vesicles as a liquid biopsy for brain insulin dysregulation in Alzheimer’s disease and related disorders. Alzheimers Dement 21, e14497 (2025). 10.1002/alz.14497

11 Kumar, A., Nader, M. A. & Deep, G. Emergence of Extracellular Vesicles as "Liquid Biopsy" for Neurological Disorders: Boom or Bust. Pharmacol Rev 76, 199–227 (2024). 10.1124/pharmrev.122.000788

12 Cleary, J. A., Kumar, A., Craft, S. & Deep, G. Neuron-derived extracellular vesicles as a liquid biopsy for brain insulin dysregulation in Alzheimer’s disease and related disorders. Alzheimers Dement, e14497 (2025). 10.1002/alz.14497

13 Kumar, A. et al. Brain cell-derived exosomes in plasma serve as neurodegeneration biomarkers in male cynomolgus monkeys self-administrating oxycodone. EBioMedicine 63, 103192 (2021). 10.1016/j.ebiom.2020.103192

14 Becker, D., Hirsch, A. G., Bender, L., Lingner, T., Salinas, G. & Krebber, H. Nuclear Pre-snRNA Export Is an Essential Quality Assurance Mechanism for Functional Spliceosomes. Cell Rep 27, 3199–3214 e3193 (2019). 10.1016/j.celrep.2019.05.031

15 Gebert, L. F. R. & MacRae, I. J. Regulation of microRNA function in animals. Nat Rev Mol Cell Biol 20, 21–37 (2019). 10.1038/s41580-018-0045-7

16 Telonis, A. G. et al. Dissecting tRNA-derived fragment complexities using personalized transcriptomes reveals novel fragment classes and unexpected dependencies. Oncotarget 6, 24797–24822 (2015). 10.18632/oncotarget.4695

17 O’Brien, J., Hayder, H., Zayed, Y. & Peng, C. Overview of MicroRNA Biogenesis, Mechanisms of Actions, and Circulation. Front Endocrinol (Lausanne) 9, 402 (2018). 10.3389/fendo.2018.00402

18 Li, M. L., Wang, W. & Jin, Z. B. Circular RNAs in the Central Nervous System. Front Mol Biosci 8, 629593 (2021). 10.3389/fmolb.2021.629593

19 Barros, F. M., Carneiro, F., Machado, J. C. & Melo, S. A. Exosomes and Immune Response in Cancer: Friends or Foes? Front Immunol 9, 730 (2018). 10.3389/fimmu.2018.00730

20 Whiteside, T. L. Exosomes and tumor-mediated immune suppression. J Clin Invest 126, 1216–1223 (2016). 10.1172/JCI81136

21 Fitz, N. F., Sahu, A., Lu, Y., Ambrosio, F., Lefterov, I. & Koldamova, R. Extracellular Vesicles in Young Serum Contribute to the Restoration of Age-Related Brain Transcriptomes and Cognition in Old Mice. Int J Mol Sci 24 (2023). 10.3390/ijms241612550

22 Cotman, C. W. & Berchtold, N. C. Physical activity and the maintenance of cognition: learning from animal models. Alzheimers Dement 3, S30–37 (2007). 10.1016/j.jalz.2007.01.013

23 Marosi, K. et al. Long-term exercise treatment reduces oxidative stress in the hippocampus of aging rats. Neuroscience 226, 21–28 (2012). 10.1016/j.neuroscience.2012.09.001

24 Steiner, J. L., Murphy, E. A., McClellan, J. L., Carmichael, M. D. & Davis, J. M. Exercise training increases mitochondrial biogenesis in the brain. J Appl Physiol (1985) 111, 1066–1071 (2011). 10.1152/japplphysiol.00343.2011

25 Delezie, J. & Handschin, C. Endocrine Crosstalk Between Skeletal Muscle and the Brain. Front Neurol 9, 698 (2018). 10.3389/fneur.2018.00698

26 Kostka, M., Morys, J., Malecki, A. & Nowacka-Chmielewska, M. Muscle-brain crosstalk mediated by exercise-induced myokines - insights from experimental studies. Front Physiol 15, 1488375 (2024). 10.3389/fphys.2024.1488375

27 Murphy, R. M., Watt, M. J. & Febbraio, M. A. Metabolic communication during exercise. Nat Metab 2, 805–816 (2020). 10.1038/s42255-020-0258-x

28 Nederveen, J. P., Warnier, G., Di Carlo, A., Nilsson, M. I. & Tarnopolsky, M. A. Extracellular Vesicles and Exosomes: Insights From Exercise Science. Front Physiol 11, 604274 (2020). 10.3389/fphys.2020.604274

29 Vechetti, I. J., Jr., Valentino, T., Mobley, C. B. & McCarthy, J. J. The role of extracellular vesicles in skeletal muscle and systematic adaptation to exercise. J Physiol 599, 845–861 (2021). 10.1113/JP278929

30 Whitham, M. et al. Extracellular Vesicles Provide a Means for Tissue Crosstalk during Exercise. Cell Metab 27, 237–251 e234 (2018). 10.1016/j.cmet.2017.12.001

31 Alvarez-Erviti, L., Seow, Y., Yin, H., Betts, C., Lakhal, S. & Wood, M. J. Delivery of siRNA to the mouse brain by systemic injection of targeted exosomes. Nat Biotechnol 29, 341–345 (2011). 10.1038/nbt.1807

32 Li, J. J. et al. In vivo evidence for the contribution of peripheral circulating inflammatory exosomes to neuroinflammation. J Neuroinflammation 15, 8 (2018). 10.1186/s12974-017-1038-8

33 Katsuda, T. et al. Human adipose tissue-derived mesenchymal stem cells secrete functional neprilysin-bound exosomes. Sci Rep 3, 1197 (2013). 10.1038/srep01197

34 Morad, G. et al. Tumor-Derived Extracellular Vesicles Breach the Intact Blood-Brain Barrier via Transcytosis. ACS Nano 13, 13853–13865 (2019). 10.1021/acsnano.9b04397

35 Jia, J., Wang, L., Zhou, Y., Zhang, P. & Chen, X. Muscle-derived extracellular vesicles mediate crosstalk between skeletal muscle and other organs. Front Physiol 15, 1501957 (2024). 10.3389/fphys.2024.1501957

36 Aswad, H. et al. Exosomes participate in the alteration of muscle homeostasis during lipid-induced insulin resistance in mice. Diabetologia 57, 2155–2164 (2014). 10.1007/s00125-014-3337-2

37 Choi, J. S. et al. Exosomes from differentiating human skeletal muscle cells trigger myogenesis of stem cells and provide biochemical cues for skeletal muscle regeneration. J Control Release 222, 107–115 (2016). 10.1016/j.jconrel.2015.12.018

38 Do, M. K. et al. Time-coordinated prevalence of extracellular HGF, FGF2 and TGF-beta3 in crush-injured skeletal muscle. Anim Sci J 83, 712–717 (2012). 10.1111/j.1740-0929.2012.01057.x

39 Hoier, B., Prats, C., Qvortrup, K., Pilegaard, H., Bangsbo, J. & Hellsten, Y. Subcellular localization and mechanism of secretion of vascular endothelial growth factor in human skeletal muscle. FASEB J 27, 3496–3504 (2013). 10.1096/fj.12-224618

40 Bean, A. C. et al. Neuromuscular electrical stimulation enhances the ability of serum extracellular vesicles to regenerate aged skeletal muscle after injury. Exp Gerontol 177, 112179 (2023). 10.1016/j.exger.2023.112179

41 Kanaya, A. M. et al. Total and regional adiposity and cognitive change in older adults: The Health, Aging and Body Composition (ABC) study. Arch Neurol 66, 329–335 (2009). 10.1001/archneurol.2008.570

42 https://healthabc.nia.nih.gov/. Introducing the Health ABC Study: The Dynamics of Health, Aging, and Body Composition.

43 Barha, C. K. et al. Sex-dependent effect of the BDNF Val66Met polymorphism on executive functioning and processing speed in older adults: evidence from the health ABC study. Neurobiol Aging 74, 161–170 (2019). 10.1016/j.neurobiolaging.2018.10.021

44 Kuller, L. H. et al. Risk factors for dementia in the cardiovascular health cognition study. Neuroepidemiology 22, 13–22 (2003). 10.1159/000067109

45 Andrews, J. S., Desai, U., Kirson, N. Y., Zichlin, M. L., Ball, D. E. & Matthews, B. R. Disease severity and minimal clinically important differences in clinical outcome assessments for Alzheimer’s disease clinical trials. Alzheimers Dement (N Y) 5, 354–363 (2019). 10.1016/j.trci.2019.06.005

46 Mishra, S. et al. A Liquid Biopsy-Based Approach to Isolate and Characterize Adipose Tissue-Derived Extracellular Vesicles from Blood. ACS Nano 17, 10252–10268 (2023). 10.1021/acsnano.3c00422

47 Isakova, A., Fehlmann, T., Keller, A. & Quake, S. R. A mouse tissue atlas of small noncoding RNA. Proc Natl Acad Sci U S A 117, 25634–25645 (2020). 10.1073/pnas.2002277117

48 Zhou, Y. et al. Metascape provides a biologist-oriented resource for the analysis of systems-level datasets. Nat Commun 10, 1523 (2019). 10.1038/s41467-019-09234-6

49 Rosano, C. et al. Association between physical and cognitive function in healthy elderly: the health, aging and body composition study. Neuroepidemiology 24, 8–14 (2005). 10.1159/000081043

50 Rosano, C., Studenski, S. A., Aizenstein, H. J., Boudreau, R. M., Longstreth, W. T., Jr. & Newman, A. B. Slower gait, slower information processing and smaller prefrontal area in older adults. Age Ageing 41, 58–64 (2012). 10.1093/ageing/afr113

51 Li, J. et al. COMPSRA: a COMprehensive Platform for Small RNA-Seq data Analysis. Sci Rep 10, 4552 (2020). 10.1038/s41598-020-61495-0

52 Qiu, W. et al. Transcriptome-wide piRNA profiling in human brains of Alzheimer’s disease. Neurobiol Aging 57, 170–177 (2017). 10.1016/j.neurobiolaging.2017.05.020

53 Roy, J., Sarkar, A., Parida, S., Ghosh, Z. & Mallick, B. Small RNA sequencing revealed dysregulated piRNAs in Alzheimer’s disease and their probable role in pathogenesis. Mol Biosyst 13, 565–576 (2017). 10.1039/c6mb00699j

54 Alsop, E. et al. A Novel Tissue Atlas and Online Tool for the Interrogation of Small RNA Expression in Human Tissues and Biofluids. Front Cell Dev Biol 10, 804164 (2022). 10.3389/fcell.2022.804164

55 Keller, A. et al. miRNATissueAtlas2: an update to the human miRNA tissue atlas. Nucleic Acids Res 50, D211–D221 (2022). 10.1093/nar/gkab808

56 Smal, M. et al. Small non-coding RNA transcriptomic profiling in adult and fetal human brain. Sci Data 11, 767 (2024). 10.1038/s41597-024-03604-6

57 Rybak-Wolf, A. et al. Circular RNAs in the Mammalian Brain Are Highly Abundant, Conserved, and Dynamically Expressed. Mol Cell 58, 870–885 (2015). 10.1016/j.molcel.2015.03.027

58 Wu, W., Ji, P. & Zhao, F. CircAtlas: an integrated resource of one million highly accurate circular RNAs from 1070 vertebrate transcriptomes. Genome Biol 21, 101 (2020). 10.1186/s13059-020-02018-y

59 Patterson, S. A., Deep, G. & Brinkley, T. E. Detection of the receptor for advanced glycation endproducts in neuronally-derived exosomes in plasma. Biochem Biophys Res Commun 500, 892–896 (2018). 10.1016/j.bbrc.2018.04.181

60 Corona, J. C. & Duchen, M. R. PPARgamma as a therapeutic target to rescue mitochondrial function in neurological disease. Free Radic Biol Med 100, 153–163 (2016). 10.1016/j.freeradbiomed.2016.06.023

61 Abramowicz, A. & Story, M. D. The Long and Short of It: The Emerging Roles of Non-Coding RNA in Small Extracellular Vesicles. Cancers (Basel) 12 (2020). 10.3390/cancers12061445

62 Chen, X. et al. Small extracellular vesicles from young plasma reverse age-related functional declines by improving mitochondrial energy metabolism. Nat Aging 4, 814–838 (2024). 10.1038/s43587-024-00612-4

63 Shin, H. E., Won, C. W. & Kim, M. Circulating small non-coding RNA profiling for identification of older adults with low muscle strength and physical performance: A preliminary study. Exp Gerontol 197, 112598 (2024). 10.1016/j.exger.2024.112598

64 Wang, Y. Q. et al. LncRNA FOXC2-AS1 regulated proliferation and apoptosis of vascular smooth muscle cell through targeting miR-1253/FOXF1 axis in atherosclerosis. Eur Rev Med Pharmacol Sci 24, 3302–3314 (2020). 10.26355/eurrev_202003_20698

65 Goodpaster, B. H. & Sparks, L. M. Metabolic Flexibility in Health and Disease. Cell Metab 25, 1027–1036 (2017). 10.1016/j.cmet.2017.04.015

66 Wang, H. et al. miR-183 and miR-96 orchestrate both glucose and fat utilization in skeletal muscle. EMBO Rep 22, e52247 (2021). 10.15252/embr.202052247

67 Sanchez-Gonzalez, C. et al. Dysfunctional oxidative phosphorylation shunts branched-chain amino acid catabolism onto lipogenesis in skeletal muscle. EMBO J 39, e103812 (2020). 10.15252/embj.2019103812

68 Koh, J. H. et al. TFAM Enhances Fat Oxidation and Attenuates High-Fat Diet-Induced Insulin Resistance in Skeletal Muscle. Diabetes 68, 1552–1564 (2019). 10.2337/db19-0088

69 Lago-Docampo, M. et al. Overexpression of miR-3168 impairs angiogenesis Pulmonary Arterial Hypertension: Insights from circulating miRNA analysis. medRxiv, 2024.2004.2030.24306656 (2024). 10.1101/2024.04.30.24306656

70 Alique, M., Bodega, G., Giannarelli, C., Carracedo, J. & Ramirez, R. MicroRNA-126 regulates Hypoxia-Inducible Factor-1alpha which inhibited migration, proliferation, and angiogenesis in replicative endothelial senescence. Sci Rep 9, 7381 (2019). 10.1038/s41598-019-43689-3

71 Pinto-Hernandez, P. et al. Training-induced plasma miR-29a-3p is secreted by skeletal muscle and contributes to metabolic adaptations to resistance exercise in mice. Mol Metab 98, 102173 (2025). 10.1016/j.molmet.2025.102173

72 Tian, Q., Lee, P. R., Walker, K. A. & Ferrucci, L. Energizing Mitochondria to Prevent Mobility Loss in Aging: Rationale and Hypotheses. Exerc Sport Sci Rev 51, 96–102 (2023). 10.1249/JES.0000000000000315

73 Rehman, S. U., Ali, R., Zhang, H., Zafar, M. H. & Wang, M. Research progress in the role and mechanism of Leucine in regulating animal growth and development. Front Physiol 14, 1252089 (2023). 10.3389/fphys.2023.1252089

74 Kocher, M. A., Huang, F. W., Le, E. & Good, D. J. Snord116 Post-transcriptionally Increases Nhlh2 mRNA Stability: Implications for Human Prader-Willi Syndrome. Hum Mol Genet 30, 1101–1110 (2021). 10.1093/hmg/ddab103

75 Joseph, A. M., Adhihetty, P. J. & Leeuwenburgh, C. Beneficial effects of exercise on age-related mitochondrial dysfunction and oxidative stress in skeletal muscle. J Physiol 594, 5105–5123 (2016). 10.1113/JP270659

76 Picca, A. et al. Mitochondrial quality control mechanisms as molecular targets in cardiac ageing. Nat Rev Cardiol 15, 543–554 (2018). 10.1038/s41569-018-0059-z

77 Amorim, J. A., Coppotelli, G., Rolo, A. P., Palmeira, C. M., Ross, J. M. & Sinclair, D. A. Mitochondrial and metabolic dysfunction in ageing and age-related diseases. Nat Rev Endocrinol 18, 243–258 (2022). 10.1038/s41574-021-00626-7

78 Luo, L. et al. Mitochondrial-related microRNAs and their roles in cellular senescence. Front Physiol 14, 1279548 (2023). 10.3389/fphys.2023.1279548

