## Supplemental Figure 1 for "Serum-derived extracellular vesicles as biological indicator of mobility resilience in older adults"

### Supplementary information

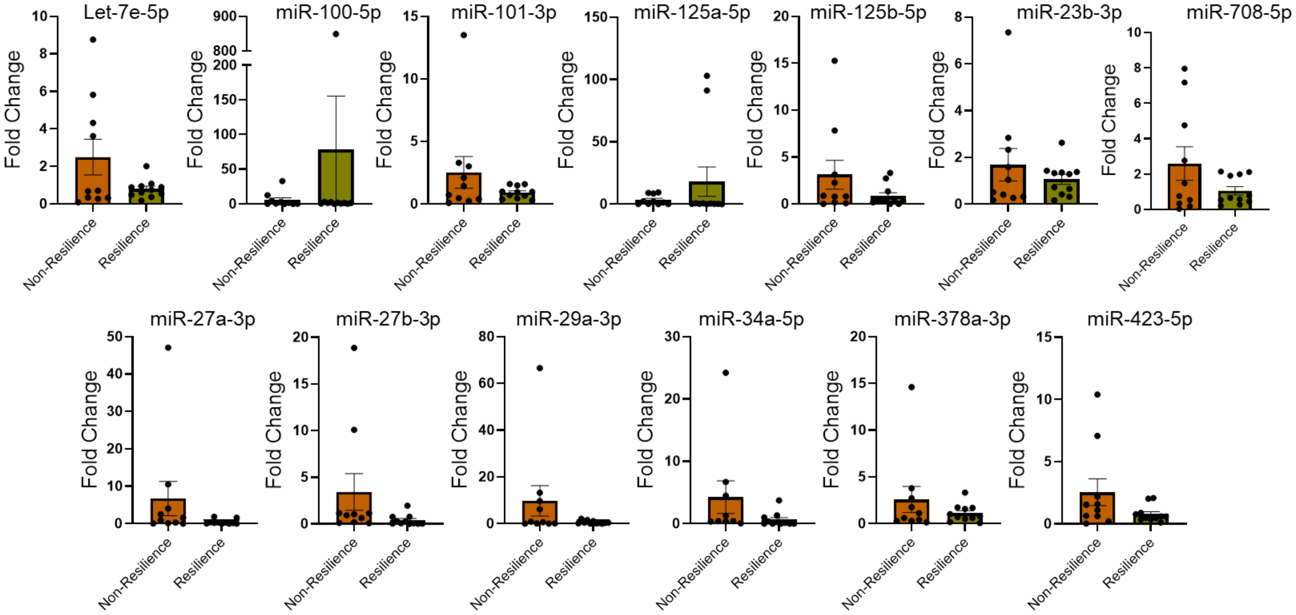


**Supplemental Figure 1.** The expression of 13 miRNAs was analyzed in MDE isolated from mobility resilience and non-resilience groups by RT-PCR. cel-miR-39 was used to normalize the expression of miRNAs as described in methods, and fold change was calculated using non-resilience group as reference. Statistical significance was determined by unpaired t-test.
